# Phase 1 Assessment of the Safety and Immunogenicity of an mRNA- Lipid Nanoparticle Vaccine Candidate Against SARS-CoV-2 in Human Volunteers

**DOI:** 10.1101/2020.11.09.20228551

**Authors:** Peter Kremsner, Philipp Mann, Jacobus Bosch, Rolf Fendel, Julian J. Gabor, Andrea Kreidenweiss, Arne Kroidl, Isabel Leroux-Roels, Geert Leroux-Roels, Christoph Schindler, Mirjam Schunk, Thirumalaisamy P. Velavan, Mariola Fotin-Mleczek, Stefan Müller, Gianluca Quintini, Oliver Schönborn-Kellenberger, Dominik Vahrenhorst, Thomas Verstraeten, Lisa Walz, Olaf-Oliver Wolz, Lidia Oostvogels

**Affiliations:** Institute of Tropical Medicine, University Hospital Tübingen, Tübingen, Germany; Ghent University Hospital, Ghent, Belgium; Division of Infectious Diseases and Tropical Medicine, University Hospital, LMU Munich, Germany; German Center for Infection Research (DZIF), partner site Munich, Germany; Hannover Medical School (MHH), Hannover, Germany; Curevac AG, Frankfurt, Germany; P95 Epidemiology and Pharmacovigilance, Leuven, Belgium; Cogitars, Heidelberg, Germany; Curevac AG, Tübingen, Germany; German Center for Infection Research (DZIF), partner site Tübingen, Tübingen, Germany; Centre de Recherches Médicales de Lambaréné, Lambaréné, Gabon

## Abstract

There is an urgent need for vaccines to counter the COVID-19 pandemic due to infections with severe acute respiratory syndrome coronavirus (SARS-CoV-2). Evidence from convalescent sera and preclinical studies has identified the viral Spike (S) protein as a key antigenic target for protective immune responses. We have applied an mRNA-based technology platform, RNActive^®^, to develop CVnCoV which contains sequence optimized mRNA coding for a stabilized form of S protein encapsulated in lipid nanoparticles (LNP). Following demonstration of protective immune responses against SARS-CoV-2 in animal models we performed a dose-escalation phase 1 study in healthy 18-60 year-old volunteers.

This interim analysis shows that two doses of CVnCoV ranging from 2 μg to 12 μg per dose, administered 28 days apart were safe. No vaccine-related serious adverse events were reported. There were dose-dependent increases in frequency and severity of solicited systemic adverse events, and to a lesser extent of local reactions, but the majority were mild or moderate and transient in duration. Immune responses when measured as IgG antibodies against S protein or its receptor-binding domain (RBD) by ELISA, and SARS-CoV-2-virus neutralizing antibodies measured by micro-neutralization, displayed dose-dependent increases. Median titers measured in these assays two weeks after the second 12 μg dose were comparable to the median titers observed in convalescent sera from COVID-19 patients. Seroconversion (defined as a 4-fold increase over baseline titer) of virus neutralizing antibodies two weeks after the second vaccination occurred in all participants who received 12 μg doses.

Preliminary results in the subset of subjects who were enrolled with known SARS-CoV-2 seropositivity at baseline show that CVnCoV is also safe and well tolerated in this population, and is able to boost the pre-existing immune response even at low dose levels.

Based on these results, the 12 μg dose is selected for further clinical investigation, including a phase 2b/3 study that will investigate the efficacy, safety, and immunogenicity of the candidate vaccine CVnCoV.

## INTRODUCTION

The global COVID-19 pandemic due to the severe acute respiratory syndrome coronavirus 2 (SARS-CoV-2) is causing uncontrolled illness around the world with almost 50 million cases and over 1 million deaths [1]. Although many infections are asymptomatic or mild, more severe cases produce respiratory distress which requires mechanical ventilation in intensive care and can result in death [2]. Increasing SARS-CoV-2 infection rates may overwhelm critical care capacity, with a consequential increase in mortality rate, highlighting the urgent need for an effective prophylactic vaccine to protect the immunologically naïve population. A major research and development effort to produce effective SAR-CoV-2 vaccines has been launched globally, with 47 candidates currently in clinical testing [3]. One new approach applied in some of these programs, is the use of mRNA coding for the required protein antigen to produce a human SARS-CoV-2 vaccine [4].

CureVac has developed and established an mRNA-based technology, RNActive^®^, for accelerated development of human vaccines [5]. Proof-of-concept was demonstrated in a first-in-human phase 1 study using chemically unmodified mRNA coding for rabies virus glycoprotein (RABV-G) [6]. That study found low responses of anti-RABV-G neutralizing antibodies that were dependent upon the route and mode of administration of the mRNA injection. Preclinical studies demonstrated improvement of the immune responses in animal models by encapsulating the mRNA in lipid nanoparticles (LNP) [7]. Another human phase 1 study was performed with RABV-G mRNA-LNP formulations which found that two 1 or 2 μg doses elicited immune responses comparable to a three-dose regimen of a licensed rabies vaccine, and with acceptable tolerability [8].

In response to the global COVID-19 pandemic, the RNActive^®^ technology platform has been applied to CVnCoV, an mRNA-LNP vaccine aimed at preventing SARS-CoV-2 infection [9]. The target antigen for many of the SARS-CoV-2 vaccines in development is the glycosylated spike (S) protein on the viral surface. S proteins from MERS [10] and SARS-CoV viruses [11] are essential for viral binding and uptake into mammalian cells, and this is now confirmed for the SARS-CoV-2 S protein [12,13]. The trimeric S protein interacts with human angiotensin-converting enzyme 2 (ACE2) receptors to allow intracellular entry of the virus following proteolytic cleavage of the S protein into its S1 and S2 domains leading to fusion with the cell membrane and release into the cell cytoplasm for viral replication [12–15]. Inhibition of S protein cleavage into the S1 and S2 domains by protease inhibitors [14], antibodies to the receptor-binding domain (RBD) of the S protein [16], or antibodies to S protein from convalescent COVID-19 patients [13] have all been protective in preclinical models, highlighting this protein as the target for vaccine development. CVnCoV consists of non-chemically modified mRNA encoding full-length S protein, encapsulated in LNP. The coded S protein includes two proline mutations (S-2P) previously been shown to stabilize the conformation of the S proteins for MERS-CoV [9] and SARS-CoV [11]. mRNA was optimized to provide a high expression level of S protein and a moderate activation of innate immunity. In rodent models, CVnCoV induces neutralizing antibodies and T cell responses. and provides lung protection in a hamster SARS-CoV-2-challenge model [9].

We report an interim analysis of the first data from an ongoing first-in-human phase 1 CVnCoV trial of a two-dose primary schedule to assess the safety, reactogenicity, and immunogenic it y in healthy adults in two age strata (18–40 years and 41–60 years). Both SARS-CoV-2 naïve and previously infected participants are included in the trial to ensure that pre-existing immunity has no effect on the assessed parameters.

## METHODS

The first-in-human, placebo-controlled, blinded phase 1 trial of CVnCoV enrolled healthy adults (18 to 60 years). This dose-finding trial is conducted in Hannover, Munich and Tübingen, Germany, and Ghent, Belgium. The study protocol was approved by the appropriate Investigational Review Boards (IRB) and national regulatory authority for each site, and was registered with ClinicalTrials.gov (Identifier: NCT 04449276). All study procedures were performed according to International Council for Harmonization of Technical Requirements for Pharmaceuticals for Human Use and Good Clinical Practice guidelines. All participants provided written informed consent at enrollment. The study was monitored for safety by an internal Safety Review Committee (iSRC) and a Data Safety Monitoring Board (DSMB) composed of independent external vaccine experts.

The primary objective was the evaluation of safety and reactogenicity of 1 or 2 doses of different dosages of CVnCoV administered by intramuscular injection 28 days apart. The main secondary objectives were the evaluation of the humoral immune response measured by SARS-CoV-2-S protein-specific IgG and RBD IgG (ELISA) antibodies, as well as SARS-CoV-2 virus neutralizing antibodies.

Since screening for SARS-CoV-2 serostatus at baseline in large clinical studies would be unpractical, a subset of SARS-CoV-2 seropositive participants were also included in the study to assess if the baseline serostatus impacts any of the parameters assessed.

### Participants

Eligible participants of either sex, in good health based on medical history and examination at screening, were enrolled in two equal age groups (18–40 and 41–60 years). Main inclusion criteria were a body mass index (BMI) ≥18.0 and ≤ 30.0kg/m^2^ and being available for the duration of the study. Main exclusion criteria were a known elevated risk of exposure to SARS-CoV-2 infection (e.g. healthcare personnel or those directly involved in patient care or long-term care), or any history of COVID-19 infection or exposure to a COVID-19 infected individual with two weeks prior to the study. Each dose level includes a subset of known SARS-CoV-2 seropositive participants for whom the earlier mentioned exclusion criteria did not apply. The following exclusion criteria were to be applied to all subjects: any current or previous history of immunosuppressive disorder or therapy, any previous confirmed infection with SARS or MERS, and any known allergy to a vaccine component. Also excluded were active smokers within the previous year, pregnant or breastfeeding women, study sponsors, and study staff employees or relatives. Women of child-bearing potential were required to have a negative pregnancy test within three days before receiving their first vaccination, and to use an approved highly effective form of contraception from one month before the first vaccination until 3 months after the last vaccination.

### Study design

This was a dosage escalation study, starting with 2 μg CVnCoV and progressively increased in subsequent groups with dosages of 4, 6, 8, and 12 μg. Higher dosages of 16 and 20 μg are currently being administered. For each dosage group there was a sentinel cohort of two participants in each of the two age groups with no history of COVID-19. Sentinels were vaccinated (two vaccinees per age group, open label) and monitored for 60 hours. The iSRC and DSMB chair assessed the safety data from the first 24 hours of these sentinels before approving the vaccination of four additional participants in each age group (open label, subjects without a history of COVID-19). After assessing safety data for 60 hours, the iSRC and DSMB approved the vaccination of the remaining participants of that dosage group (including placebo subjects and subjects known to be seropositive for SARS-CoV-2, randomized and blinded) and the sentinels of the next higher dosage group. This procedure was then repeated until all groups were vaccinated for dose levels up to 8 µg. The procedure was the same for 12 µg dose, but all subjects were open label, without placebo control.

### Vaccine

The study vaccine, CVnCoV, is an investigational LNP-formulated RNActive^®^ SARS-CoV-2 vaccine composed of the active pharmaceutical ingredient, an mRNA that encodes a pre-fusion conformation stabilized version of the full length spike (S) protein of SARS-CoV-2 virus, and four lipid components: cholesterol, 1,2-distearoyl-sn-glycero-3-phosphocholine (DSPC), PEG-ylated lipid and a cationic lipid. The placebo administered in the control arms consists of 0.9% NaCl. Each 0.5 mL dose was administered by intramuscular injection in the deltoid. All administrations were performed by unblinded study staff who had no role in data collection for safety or immunogenicity assessments.

### Safety assessments

All participants remained under direct supervision of site personnel for 4 hours following administration of their assigned injection. Participants then recorded in diary cards solicited local (injection-site pain, redness, swelling, and itching) and systemic (headache, fatigue, chills, myalgia, arthralgia, nausea/vomiting, and diarrhea) adverse events and daily temperature using a supplied thermometer for 7 days after each vaccination, and any unsolicited adverse events until the next study visit 28 days after vaccination. All solicited AEs were graded for severity as Grade 1 (mild: easily tolerated, causing minimal discomfort and not interfering with everyday activities), Grade 2 (moderate: causes sufficient discomfort to interfere with normal everyday activities) and Grade 3 (severe: prevents normal everyday activities) using the FDA Grading Scale [17] (see Supplementary material). The investigator reviewed the severity gradings on the diary cards and used their clinical judgement to assess causality as either related (there is a reasonable causal relationship between the trial vaccine and the AE) or unrelated (there is no reasonable causal relationship between the trial vaccine and the AE). Blood samples were also drawn on Days 1, 2, 8, 30 and 36 to perform laboratory safety assessments, graded according to the FDA Grading Scale [17].

Serious adverse events (SAE), defined as life-threatening events, events resulting in death, events requiring inpatient hospitalization or prolongation of existing hospitalization, or events resulting in persistent disability/incapacity, were to be reported to the investigator immediately, who notified the sponsor. Investigators could also consider additional medical events to be SAEs. Monitored adverse events of special interest (AESI) included potential immune mediated diseases and COVID-19 disease. In the event of a confirmed COVID-19 infection the participant or treating health care provider were to complete a specific diary card. Safety will continue to be monitored for one year after the last vaccination.

### Immunogenicity assessments - IgG ELISA

Blood samples were drawn on Days 1 and 29, before each of the two vaccinations, and on Days 8, 15, 36, 43 and 57 for immunogenicity assessments. Sera were prepared and stored at -80°C before shipping on dry-ice for measurement of the immune responses in accordance with EMA “Guideline on bioanalytical method validation” at Vismederi S.r.l., Siena, Italy. Anti-SARS-CoV-2-specific IgG levels were measured by ELISA. Briefly, plates were coated with 1µg/ml of SARS-CoV-2 Spike (Spike S1+S2 ECD-His Recombinant Protein, Cat: 40589-V08B1; Sino Biological, Chesterbrook, PA, USA) or Spike RBD (Spike RBD-His Recombinant Protein, Cat. 40592-V08H; Sino Biological) recombinant protein. Blocking was performed in 5% milk. Coated plates were incubated with heat-inactivated (56°C for 30 min) human serum in a 1:2-fold serial dilution (starting at 1:100). Antigen-specific IgG detection was performed with goat anti-human IgG-HRP conjugate (Cat: A80-104P-93, Bethyl Laboratories, Montgomery, Texas, USA) and tetramethyl benzidine (TMB) substrate (Bethyl Laboratories, Montgomery, Texas, USA) at OD 450 nm. The titer was determined as the reciprocal of the highest serum dilution that is over the pre-determined cut-off OD value (limit of detection plus matrix effect) and reported as geometric mean titer (GMT) of duplicates. If no antibody was detectable (all dilutions below cut-off OD), an arbitrary titer value of 50 (half of the limit of quantification) was reported.

### Immunogenicity assessments – Neutralizing activity

SARS-CoV-2 virus neutralization titers were determined by a micro-neutralization assay with Cytopathic Effect (CPE)-read out as described previously [17]. In brief, heat-inactivated (56°C for 30 min) human serum was serially diluted 1:2 (starting at 1:10) and incubated with wild type SARS-CoV-2 virus strain 2019-nCov/Italy-INMI1 at 37°C 5% CO_2_ for 1 hour. Afterwards, semi-confluent Vero E6 cells (ATCC, Cat.1586) were incubated with the virus - serum mixtures at 37°C 5% CO_2_ for 3 days. Cells were assessed for virus-induced CPE by light microscopy. The neutralization titer was determined as the reciprocal of the highest serum dilution that protected more than the 50% of cells from CPE and reported as geometric mean titer (GMT) of duplicates. If no neutralization was observed, an arbitrary titer value of 5 (half of limit of quantification) was reported.

### Reference human convalescent sera

The pool of human COVID-19 convalescent sera consisted of 67 samples collected mainly between 4 to 8 weeks after diagnostic confirmation of SARS-CoV-2 infection. Sera were either purchased from MTG Group (Van Nuys, California, USA) under the MTG Group PROTOCOL NO: MTG-022, Ethic Approval Sterling Institutional Review Board ID: 3764, or were donated by the Universitätsklinikum, Tübingen. These samples included 16 sera from hospitalized patients, the remaining 51 being from patients who were not hospitalized but manifested with clear COVID-19 illness with various symptoms.

### Statistics

As this is an exploratory study results are presented descriptively. The sample size was not based on any hypothesis but was intended to allow estimation of the probability that the true rate of adverse reactions for each dose lies in an acceptable safety range. A minimum of 12 evaluable participants per dose and per age group was considered adequate for this purpose, but in anticipation of drop-outs and with uncertainty about the proportion which would subsequently be found to have prior asymptomatic exposure to SARS-CoV-2 a conservative number of 48 per group (24 in each age group) was chosen. As this is a dose-escalation study not all safety and immunogenicity are yet available for the later groups (higher doses) in this interim analysis, and safety and immunogenicity data are not presented according to age group or retrospectively confirmed baseline serostatus for SARS-CoV-2. Such analyses will be performed and full details according to these factors will be published when the full data set is available.

Safety data was analyzed in the Safety Set composed of all those who received one study administration (vaccine or placebo) and for whom any post-vaccination safety data was available. Safety data is presented descriptively as numbers of participants and percentages of each group having a specific solicited AE, together with severity. SAEs and AESIs are described by case. The primary endpoints for the safety objective were the frequencies of SAEs, Grade 3 solicited AEs within 60 hours of vaccination, frequencies and severity of solicited AEs within 7 days of vaccination, and the occurrence, intensities, and causality of unsolicited AEs with 28 days of vaccination. In this interim report of safety data we distinguis h between dosage groups, but not between age groups.

Immunogenicity was analyzed in all subjects who received both doses and who had no protocol deviation. Secondary immunogenicity endpoints are the proportions of participants seroconverting for SARS-CoV-2 S protein or RBD IgG and neutralizing antibodies, as measured by ELISA and micro-neutralization assay, respectively. Seroconversion is defined as a four-fold increase in titer over baseline. Data are presented as group median titers (with 2th and 75th percentiles) of the individual antibody GMTs. Immunogenicity data are presented graphically for individuals who received both vaccinations and per groups at time-points for which data is available.

Data are presented separately for subjects known to be SARS-CoV-2 seropositive at baseline, and who were enrolled in the 2 or 4 µg dose levels.

To analyze translation of binding into neutralizing antibodies, for every subject and visit, the ratios between VNT to ELISA RBD and ELISA Spike were generated individually, and the median drawn on those aggregations per dose and visit, and compared to the results from the sera conversion panel used.

## RESULTS

### Demographics

By the time of this interim analysis 248 adults were enrolled and assigned to the different study groups. Of these, 245 received their first vaccination or placebo injection (**Figure 1**). Compliance was good, with 231 (94%) receiving their second administration. The mean age overall was 38.6 years (± 12.9 S.D.) and there were 142 (57%) men and 106 (43%) women, the mean BMI was 23.8 kg/m^2^ (± 2.63 S.D.). The majority were described as white (237 [96%]) and non-Hispanic or Latino (242 [98%]). These demographics were consistent across the vaccine and placebo groups (**Table 1**).

**Table 1.** Demographics of the enrolled study population included in this interim analysis by group

**Tables and Figures pertaining to subjects not known to be seropositive at baseline (Table 1 to 5 and Figures 1 to 5)**
|  |  |  |  |  |  |  |  |
| --- | --- | --- | --- | --- | --- | --- | --- |
| **Group** |  | **1** | **2** | **3** | **4** | **5** | **6** |
|  |  | **2 µg** | **4 µg** | **6 µg** | **8 µg** | **12 µg** | **Placebo** |
| N= |  | 47 | 48 | 48 | 45 | 28 | 32 |
| **Age**<br>(years) | Mean | 38.2 | 39.1 | 38.6 | 38.2 | 37.4 | 40.1 |
|  | SD | 12.5 | 13.2 | 12.7 | 13.1 | 13.5 | 13.5 |
|  | range | (18–60) | (19–59) | (20–59) | (20–59) | (19–59) | (19–60) |
| **Male** | n (%) | 27 (57) | 25 (52) | 31 (65) | 27 (60) | 17 (61) | 15 (47) |
| **Female** |  | 20 (43) | 23 (48) | 17 (35) | 18 (40) | 11 (39) | 17 (53) |
| **BMI**<br>(kg/m<sup>2</sup>) | Mean | 23.6 | 24.2 | 24.3 | 23.6 | 23.6 | 23.1 |
|  | SD | (2.54) | (2.77) | (2.67) | (2.65) | (2.56) | (2.48) |
| **Race** | n (%) |  |  |  |  |  |  |
| Asian |  | 0 | 0 | 2 (4) | 2 (4) | 0 | 1 (3) |
| Brazilian |  | 1 (2) | 0 | 0 | 0 | 0 | 0 |
| Black |  | 1 (2) | 0 | 0 | 0 | 0 | 0 |
| White |  | 43 (91) | 48 (100) | 44 (92) | 43 (96) | 28 (100) | 31 (97) |
| Other or unknown |  | 1 (2) | 0 | 2 (4) | 0 | 0 | 0 |
| **Ethnicity** | n (%) |  |  |  |  |  |  |
| Hispanic or Latino |  | 3 (6) | 0 | 2 (4) | 0 | 0 | 0 |
| Not Hispanic or Latino |  | 44 (94) | 48 (100) | 46 (96) | 45 (100) | 27 (96) | 32 (100) |
| Other or unknown |  | 0 | 0 | 0 | 0 | 1 (4) | 0 |

**Figure 1.**
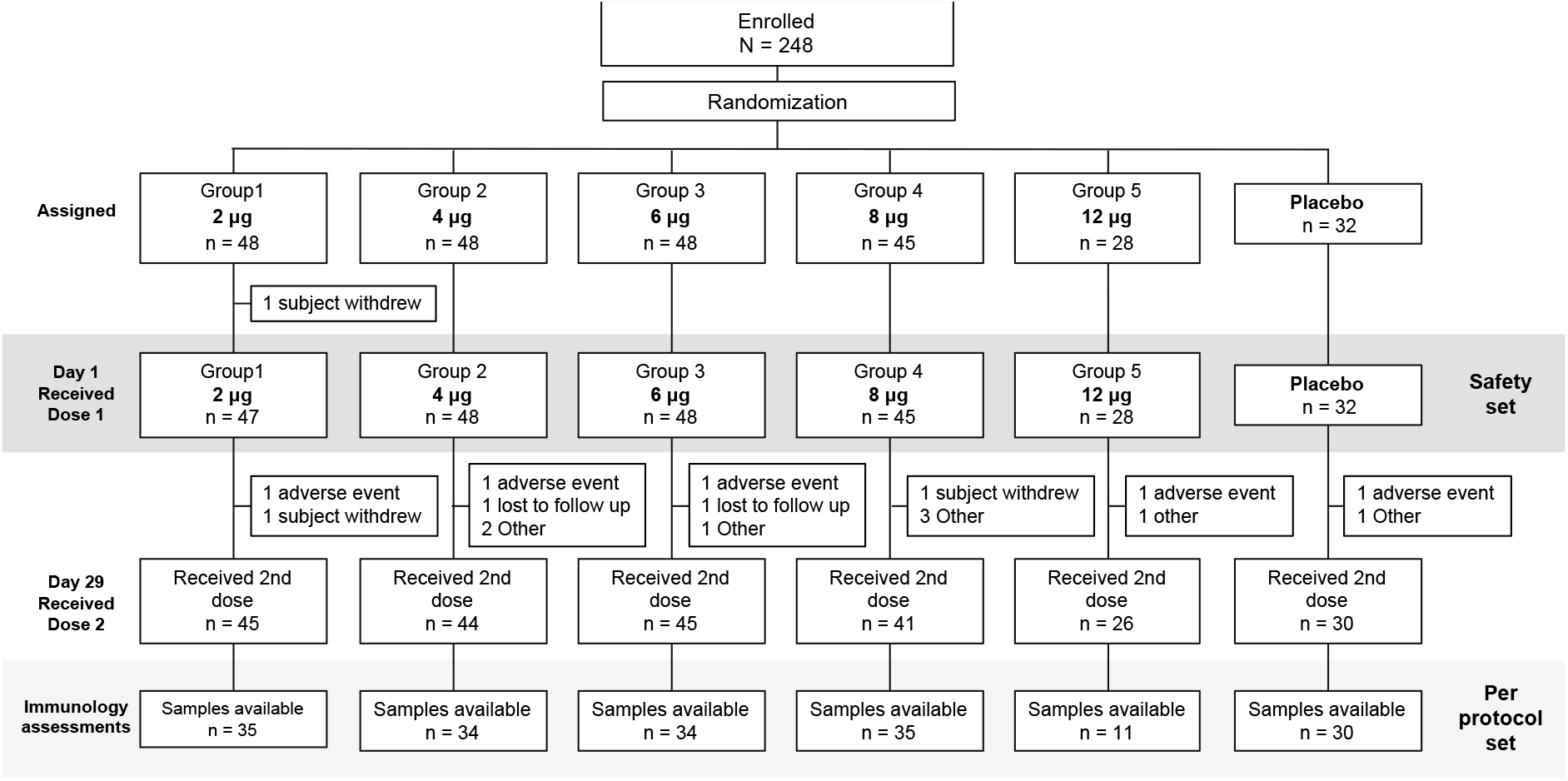
CONSORT Flow Chart

A total of 17 participants, 15 vaccinees and 2 placebo recipients, did not receive the second dose administration. Nine were unable to attend the visit, four of them because of an unrelated concurrent AE. Four vaccinees discontinued participation in the study before Day 29. Four vaccinees did not receive their second dose because of an AE following the first dose administration, three because Grade 3 AEs, and one because of a potential allergic reaction. Demography of subjects known to be SARS-CoV-2 seropositive at baseline is presented in **Table 7**.

### Safety

The primary objective to demonstrate the safety of CVnCoV was shown with no vaccine-related SAEs or AESIs reported. Three SAEs (**Table 2**) reported were cases of a complicated fracture of the humerus, abdominal pain, and monoplegia of the right foot, none of which were considered to be related to vaccination.

**Table 2.** Unsolicited AEs, SAEs and AESIs after vaccination

| Table 2. Unsolicited AEs, SAEs and AESIs after vaccination |  |  |  |  |  |  |  |
| --- | --- | --- | --- | --- | --- | --- | --- |
|  | Severity | 2 µg | 4 µg | 6 µg | 8 µg | 12 µg | Placebo |
|  | N = | 47 | 48 | 48 | 45 | 28 | 32 |
| After first dose |  |  |  |  |  |  |  |
| Unsolicited | Any | 25 (54.3) | 32 (66.7) | 35 (72.9) | 30 (66.7) | 21 (75.0) | 18 (56.3) |
|  | Related | 8 (17.4) | 19 (39.6) | 18 (37.5) | 19 (42.2) | 12 (42.9) | 5 (15.6) |
| SAEs | Any | 0 | 0 | 1 (2.1) | 0 | 1 (3.6) | 1 (3.1) |
|  | Related | 0 | 0 | 0 | 0 | 0 | 0 |
| AESI | Any | 0 | 0 | 0 | 0 | 0 | 0 |
| AEs leading to discontinuation or withdrawal | Any | 1 (2.1) | 1 (2.1) | 1 (2.1) | 0 | 1 (3.6) | 1 (3.1) |

Overall, there was a dose-dependent increase in the incidence and severity of local solicited AEs, illustrated in **Figure 2**. The vast majority of these reports were of Grade 1 and 2 injection site pain (**Table 3**); cases of severe pain usually had onset within 24 hours of vaccination, decreased in severity and resolved within 48 hours. The incidence of reactions was similar after the second dose, but lower in severity.

**Table 3.** Solicited local AEs with severity after one dose

| Table 3. Solicited local AEs with severity after one dose |  |  |  |  |  |  |  |
| --- | --- | --- | --- | --- | --- | --- | --- |
|  | Severity | 2 µg | 4 µg | 6 µg | 8 µg | 12 µg | Placebo |
|  | N = | 46 | 48 | 48 | 45 | 28 | 32 |
| After first dose |  |  |  |  |  |  |  |
| Pain | Any | 32 (69.6) | 42 (87.5) | 38 (79.2) | 40 (88.9) | 26 (92.9) | 4 (12.5) |
|  | Mild | 30 (65.2) | 38 (79.2) | 32 (66.7) | 26 (57.8) | 17 (60.7) | 4 (12.5) |
|  | Moderate | 2 (4.3) | 3 (6.3) | 5 (10.4) | 11 (24.4) | 9 (32.1) | 0 |
|  | Severe | 0 | 1 (2.1) | 1 (2.1) | 1 (2.2) | 0 | 0 |
| Redness | Any | 0 | 0 | 0 | 0 | 0 | 0 |
| Swelling | Any | 0 | 4 (8.3) | 1 (2.1) | 0 | 1 (3.6) | 0 |
|  | Mild | 0 | 4 (8.3) | 1 (2.1) | 0 | 1 (3.6) | 0 |
|  | Moderate | 0 | 0 | 0 | 0 | 0 | 0 |
|  | Severe | 0 | 0 | 0 | 0 | 0 | 0 |
| Itching | Any | 0 | 1 (2.1) | 5 (10.4) | 3 (6.7) | 2 (7.1) | 1 (3.1) |
|  | Mild | 0 | 1 (2.1) | 5 (10.4) | 3 (6.7) | 2 (7.1) | 1 (3.1) |
|  | Moderate | 0 | 0 | 0 | 0 | 0 | 0 |
|  | Severe | 0 | 0 | 0 | 0 | 0 | 0 |
| After second dose |  |  |  |  |  |  |  |
|  | N= | 44 | 44 | 45 | 41 | 26 | 30 |
| Pain | Any | 24 (54.5) | 34 (77.3) | 30 (66.7) | 34 (82.9) | 22 (84.6) | 1 (3.3) |
|  | Mild | 24 (54.5) | 32 (72.7) | 25 (55.5) | 29 (70.7) | 17 (65.4) | 1 (3.3) |
|  | Moderate | 0 | 2 (4.5) | 5 (11.1) | 5 (12.2) | 5 (19.2) | 0 |
|  | Severe | 0 | 0 | 0 | 0 | 0 | 0 |
| Redness | Any | 0 | 0 | 0 | 0 | 0 | 0 |
| Swelling | Any | 0 | 1 (2.3) | 0 | 0 | 0 | 0 |
|  | Mild | 0 | 1 (2.3) | 0 | 0 | 0 | 0 |
|  | Moderate | 0 | 0 | 0 | 0 | 0 | 0 |
|  | Severe | 0 | 0 | 0 | 0 | 0 | 0 |
| Itching | Any | 0 | 1 (2.3) | 2 (4.4) | 1 (4.9) | 1 (3.8) | 0 |
|  | Mild | 0 | 1 (2.3) | 2 (4.4) | (4.9) | 1 (3.8) | 0 |
|  | Moderate | 0 | 0 | 0 | 0 | 0 | 0 |
|  | Severe | 0 | 0 | 0 | 0 | 0 | 0 |

**Table 4a.** Solicited systemic AEs with severity within 7 days after the first dose

|  | Severity | 2 µg | 4 µg | 6 µg | 8 µg | 12 µg | Placebo |
| --- | --- | --- | --- | --- | --- | --- | --- |
|  | N = | 46 | 48 | 48 | 45 | 28 | 32 |
| **Fever** | **Any** | **3 (6.5)** | **10 (20.8)** | **16 (33.3)** | **19 (42.2)** | **16 (57.1)** | **0** |
|  | Mild | 2 (4.3) | 6 (12.5) | 11 (22.9) | 11 (24.4) | 6 (21.4) | 0 |
|  | Moderate | 1 (2.2) | 2 (4.2) | 4 (8.3) | 4 (8.9) | 6 (21.4) | 0 |
|  | Severe | 0 | 2 (4.2) | 1 (2.1) | 4 (8.9) | 4 (14.3) | 0 |
| **Headache** | **Any** | **22 (47.8)** | **36 (75.0)** | **34 (70.8)** | **38 (84.4)** | **25 (89.3)** | **10 (31.3)** |
|  | Mild | 16 (34.8) | 22 (45.8) | 15 (31.3) | 13 (28.9) | 2 (7.1) | 8 (25.0) |
|  | Moderate | 4 (8.7) | 12 (25.0) | 14 (29.2) | 19 (42.2) | 20 (71.4) | 2 (6.3) |
|  | Severe | 2 (4.3) | 2 (4.2) | 5 (10.4) | 6 (13.3) | 3 (10.7) | 0 |
| **Fatigue** | **Any** | **22 (47.8)** | **35 (72.9)** | **39 (81.3)** | **39 (86.7)** | **27 (96.4)** | **15 (46.9)** |
|  | Mild | 16 (34.8) | 20 (41.7) | 20 (41.7) | 19 (42.2) | 7 (25.0) | 13 (40.6) |
|  | Moderate | 4 (8.7) | 6 (12.5) | 14 (29.2) | 14 (31.1) | 17 (60.7) | 2 (6.3) |
|  | Severe | 2 (4.3) | 9 (18.8) | 5 (10.4) | 6 (13.3) | 3 (10.7) | 0 |
| **Chills** | **Any** | **6 (13.0)** | **15 (31.3)** | **12 (25.0)** | **20 (44.4)** | **24 (85.7)** | **0** |
|  | Mild | 3 (6.5) | 9 (18.8) | 11 (22.9) | 12 (26.7) | 6 (21.4) | 0 |
|  | Moderate | 2 (4.3) | 4 (8.3) | 8 (16.7) | 4 (8.9) | 14 (50.0) | 0 |
|  | Severe | 1 (2.2) | 2 (4.2) | 1 (2.1) | 4 (8.9) | 4 (14.3) | 0 |
| **Myalgia** | **Any** | **11 (23.9)** | **23 (47.9)** | **32 (66.7)** | **28 (62.2)** | **22 (78.6)** | **6 (18.8)** |
|  | Mild | 8 (17.4) | 14 (29.2) | 21 (43.8) | 14 (31.1) | 9 (32.1) | 6 (18.8) |
|  | Moderate | 3 (6.5) | 7 (14.6) | 10 (20.8) | 10 (22.2) | 12 (42.9) | 0 |
|  | Severe | 0 | 2 (4.2) | 1 (2.1) | 4 (8.9) | 1 (3.6) | 0 |
| **Arthralgia** | **Any** | **6 (13.0)** | **14 (29.2)** | **18 (37.5)** | **17 (37.8)** | **16 (57.1)** | **0** |
|  | Mild | 6 (13.0) | 11 (22.9) | 10 (20.8) | 10 (22.2) | 11 (39.3) | 0 |
|  | Moderate | 0 | 2 (4.2) | 6 (12.5) | 5 (11.1) | 3 (10.7) | 0 |
|  | Severe | 0 | 1 (2.1) | 2 (4.2) | 2 (4.4) | 2 (7.1) | 0 |
| **Nausea/**<br>**Vomiting** | **Any** | **3 (6.5)** | **9 (18.8)** | **9 (18.8)** | **10 (22.2)** | **11 (39.3)** | **2 (6.3)** |
|  | Mild | 2 (4.3) | 7 (14.6) | 7 (14.6) | 10 (22.2) | 7 (25.0) | 2 (6.3) |
|  | Moderate | 1 (2.2) | 2 (4.2) | 1 (2.1) | 0 | 3 (10.7) | 0 |
|  | Severe | 0 | 0 | 1 (2.1) | 0 | 1 (3.6) | 0 |
| **Diarrhea** | **Any** | **3 (6.5)** | **7 (14.6)** | **10 (20.8)** | **3 (6.7)** | **5 (17.9)** | **5 (15.6)** |
|  | Mild | 3 (6.5) | 7 (14.6) | 7 (14.6) | 3 (6.7) | 5 (17.9) | 5 (15.6) |
|  | Moderate | 0 | 0 | 3 (6.3) | 0 | 0 | 0 |
|  | Severe | 0 | 0 | 0 | 0 | 0 | 0 |

**Table 4b.** Solicited systemic AEs with severity within 7 days after the second dose

|  | Severity | 2 µg | 4 µg | 6 µg | 8 µg | 12 µg | Placebo |
| --- | --- | --- | --- | --- | --- | --- | --- |
|  | N = | 44 | 44 | 45 | 41 | 26 | 30 |
| **Fever** | **Any** | **1 (2.3)** | **3 (4.8)** | **12 (26.7)** | **12 (29.3)** | **14 (53.8)** | **0** |
|  | Mild | 1 (2.3) | 2 (4.5) | 8 (17.8) | 6 (14.6) | 5 (19.2) | 0 |
|  | Moderate | 0 | 0 | 3 (6.7) | 3 (7.3) | 6 (23.1) | 0 |
|  | Severe | 0 | 1 (2.3) | 1 (2.2) | 3 (7.2) | 3 (11.5) | 0 |
| **Headache** | **Any** | **12 (27.3)** | **26 (59.1)** | **30 (66.6)** | **33 (80.5)** | **22 (84.6)** | **7 (23.3)** |
|  | Mild | 8 (18.2) | 16 (36.4) | 17 (37.8) | 16 (39.0) | 5 (19.2) | 6 (20.0) |
|  | Moderate | 3 (6.8) | 8 (18.2) | 9 (20.0) | 12 (29.3) | 14 (53.8) | 1 (3.3) |
|  | Severe | 0 | 2 (4.5) | 4 (8.8) | 5 (12.2) | 3 (11.5) | 0 |
| **Fatigue** | **Any** | **13 (29.5)** | **25 (56.8)** | **28 (62.2)** | **34 (82.9)** | **25 (96.2)** | **5 (16.7)** |
|  | Mild | 9 (20.5) | 16 (36.3) | 16 (35.6) | 19 (46.3) | 8 (30.8) | 5 (16.7) |
|  | Moderate | 4 (9.0) | 4 (9.0) | 8 (17.8) | 10 (24.4) | 15 (57.7) | 0 |
|  | Severe | 0 | 5 (11.4) | 4 (8.9) | 5 (12.2) | 2 (7.7) | 0 |
| **Chills** | **Any** | **4 (9.1)** | **7 (15.9)** | **15 (33.3)** | **15 (36.6)** | **21 (80.8)** | **0** |
|  | Mild | 3 (6.8) | 3 (6.8) | 9 (20.0) | 8 (19.5) | 5 (19.2) | 0 |
|  | Moderate | 1 (2.3) | 3 (6.8) | 5 (11.1) | 4 (9.8) | 12 (46.2) | 0 |
|  | Severe | 0 | 1 (2.3) | 1 (2.2) | 3 (7.3) | 4 (15.4) | 0 |
| **Myalgia** | **Any** | **7 (15.9)** | **15 (34.1)** | **23 (51.1)** | **20 (48.8)** | **19 (73.1)** | **1 (3.3)** |
|  | Mild | 6 (13.3) | 9 (20.5) | 15 (33.3) | 11 (26.8) | 10 (38.5) | 1 (3.3) |
|  | Moderate | 1 (2.3) | 4 (9.1) | 7 (15.6) | 7 (17.1) | 8 (30.8) | 0 |
|  | Severe | 0 | 2 (4.5) | 1 (2.2) | 2 (4.9) | 1 (3.8) | 0 |
| **Arthralgia** | **Any** | **1 (2.3)** | **8 (18.2)** | **15 (33.3)** | **12 (29.3)** | **11 (42.3)** | **0** |
|  | Mild | 1 (2.3) | 6 (13.3) | 7 (15.5) | 9 (22.0) | 7 (26.9) | 0 |
|  | Moderate | 0 | 1 (2.3) | 4 (8.9) | 2 (4.9) | 2 (7.7) | 0 |
|  | Severe | 0 | 1 (2.3) | 2 (4.4) | 1 (2.4) | 2 (7.7) | 0 |
| **Nausea/**<br>**Vomiting** | **Any** | **1 (2.3)** | **6 (13.3)** | **6 (13.3)** | **8 (19.5)** | **7 (26.9)** | **1 (3.3)** |
|  | Mild | 1 (2.3) | 4 (9.0) | 4 (8.9) | 8 (19.5) | 5 (19.2) | 1 (3.3) |
|  | Moderate | 0 | 2 (4.5) | 1 (2.2) | 0 | 2 (7.7) | 0 |
|  | Severe | 0 | 0 | 1 (2.2) | 0 | 0 | 0 |
| **Diarrhea** | **Any** | **0** | **2 (4.5)** | **5 (11.1)** | **3 (2.4)** | **3 (11.5)** | **5 (16.7)** |
|  | Mild | 0 | 2 (4.5) | 3 (6.7) | 1 (2.4) | 3 (11.5) | 5 (16.7) |
|  | Moderate | 0 | 0 | 2 (4.4) | 0 | 0 | 0 |
|  | Severe | 0 | 0 | 0 | 0 | 0 | 0 |

**Figure 2.**
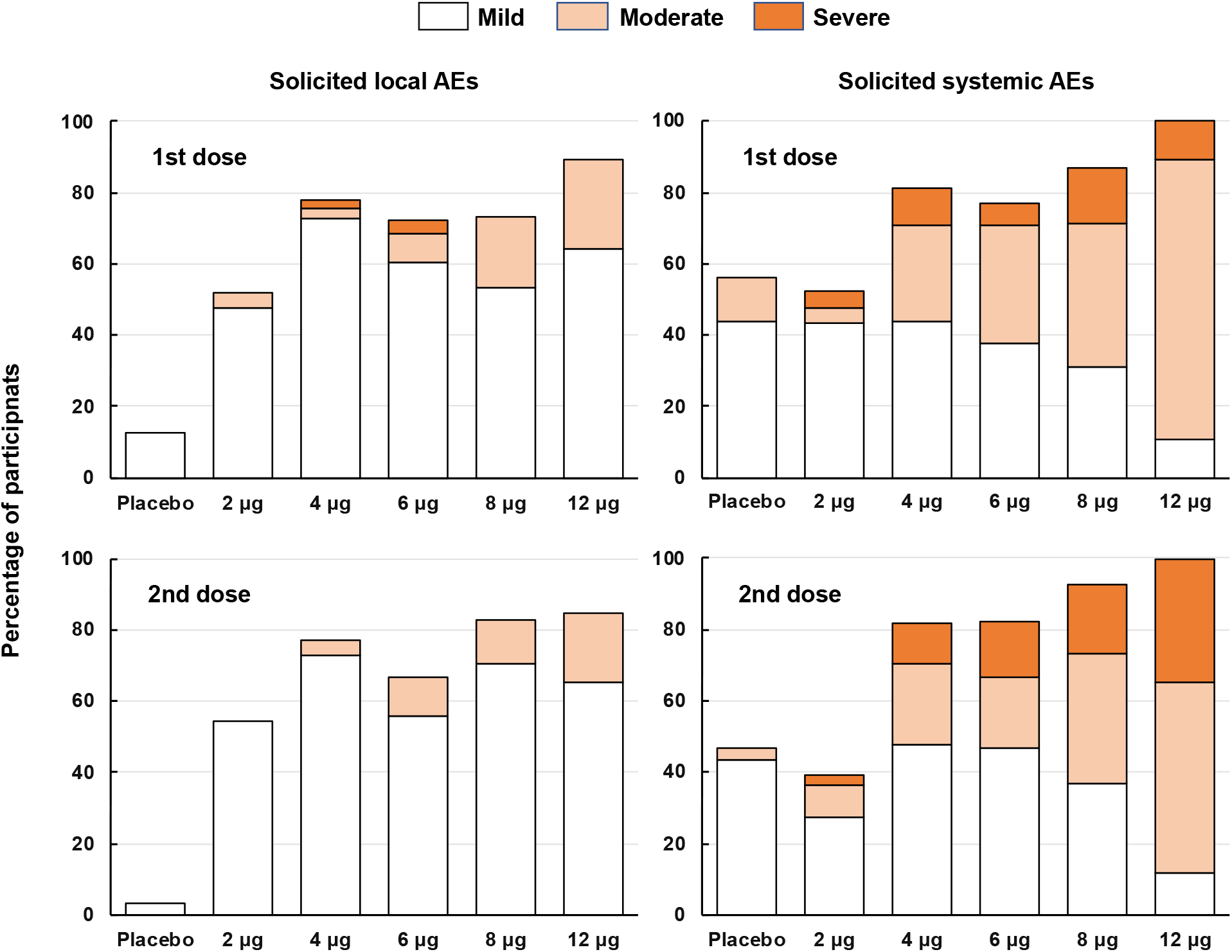
Overall incidence rates (%) of solicited local and systemic AEs per group with severity after the first and second doses

Systemic reactogenicity in terms of frequency and severity increased with dose level. All 12 μg dose participants reported at least one solicited systemic AE (**Figure 2**). Systemic AEs displayed similar overall rates after the first and second vaccinations, but the severity of these increased for the second dose in the groups receiving the 4 μg to 12 μg dose. For example, one of the 12 μg recipients had Grade 3 systemic AEs after the first dose compared with 3 after the second dose. Most Grade 3 systemic AEs had decreased in severity or resolved within 24 hours, all did so with 72 hours. The most frequent solicited systemic AEs were mild or moderate headache and fatigue, followed by myalgia and chills. Overall, fever was observed less frequently.

Unsolicited AEs were reported by the majority of participants in all groups, about half of these being considered to be related to the study procedures. Many of the unsolicited AEs were the same type as solicited AEs such as headache or fatigue but occurred outside of the 7-day solicitation period.

Laboratory adverse events were rare, with no specific pattern observed other than strictly transient lymphopenia, observed the day after vaccination and thought to represent lymphocyte redistribution related to the mode of action (data not shown) [19].

Subjects known to be SARS-CoV-2 seropositive at baseline overall experienced less reactogenicity after vaccination with either 2µg or 4µg of CVnCoV (**Tables 8 and 9**).

### Immunogenicity

Strong immune responses were observed in all vaccine groups as illustrated in Figures 3, 4 and 5 using all three assays, including in subjects who were known to be SARS-CoV-2 seropositive at baseline. As no responses were observed in placebo recipients, they are not included in the following descriptions.

**Figure 3.**
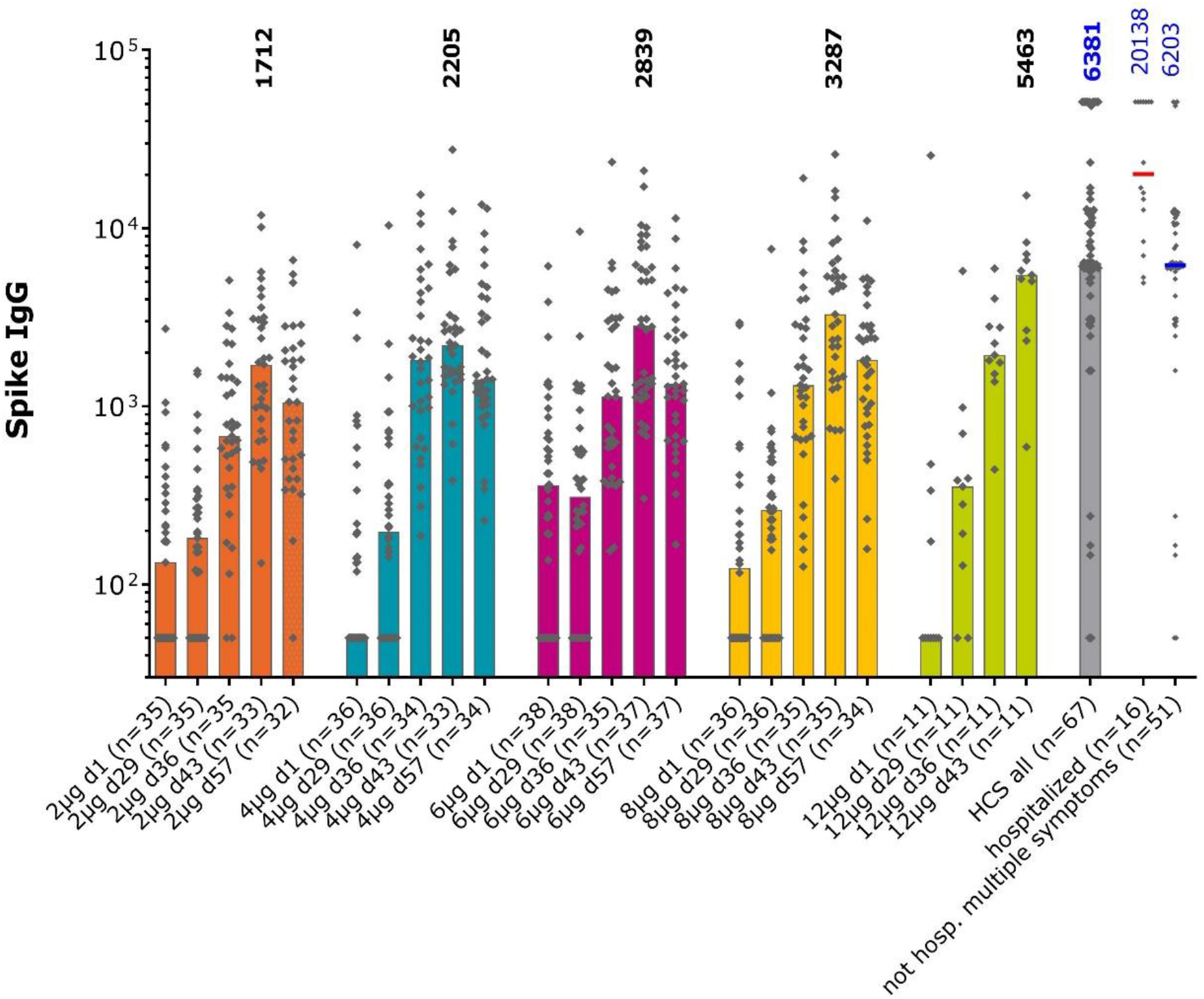
Anti-Spike protein IgG in the different study groups and convalescent sera measured by ELISA. Bars show median values per group at each study timepoint, individual GMT values for each sample shown as diamonds. Numbers show median values at Day 43, two weeks after the second vaccination, for each group and in the convalescent sera.

**Figure 4.**
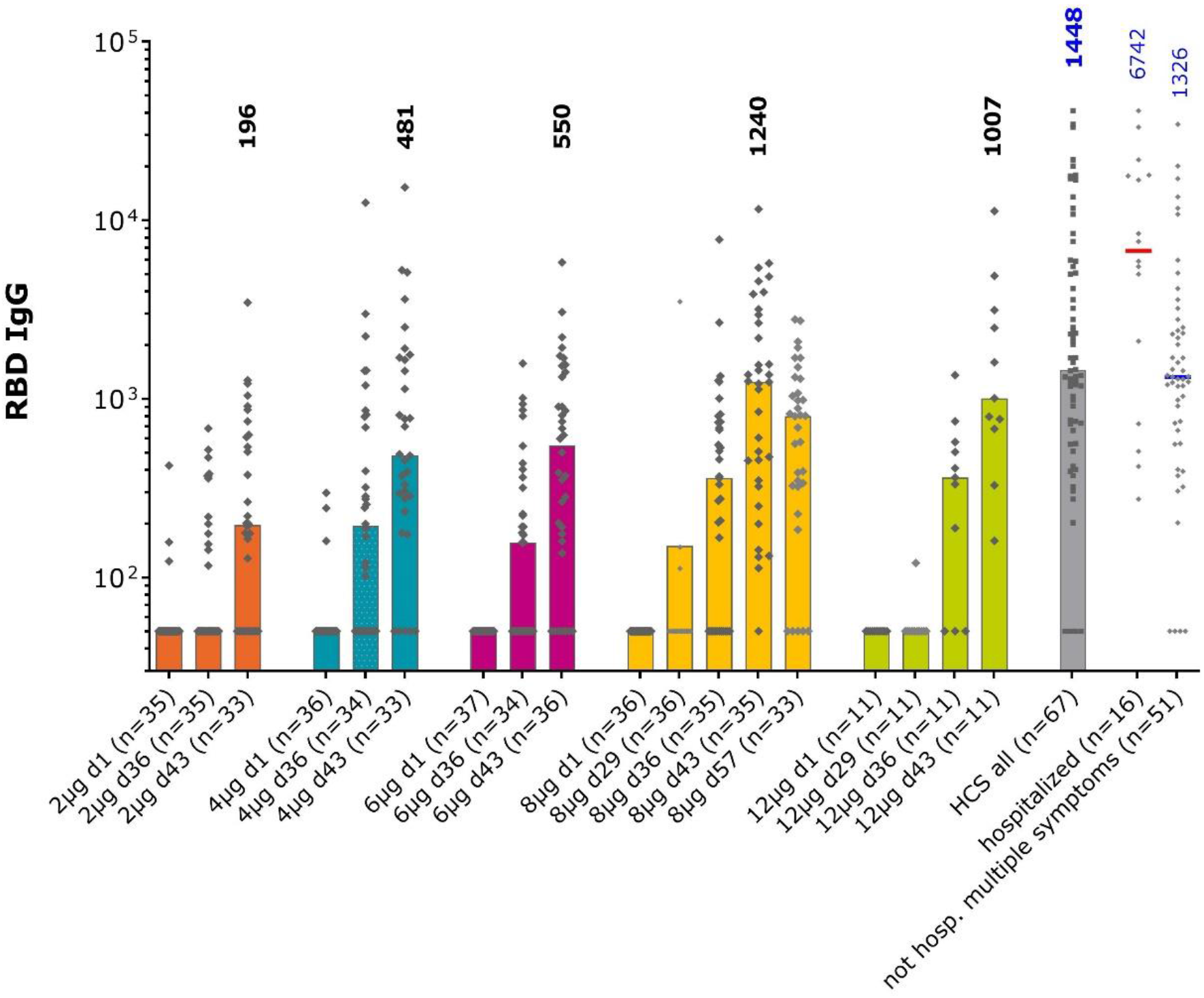
Anti-RBD IgG in the different study groups and convalescent sera measured by ELISA. Bars show median values per group at each study timepoint, individual GMT values for each sample shown as diamonds. Numbers show median values at Day 43, two weeks after the second vaccination, for each group and in the convalescent sera.

**Figure 5.**
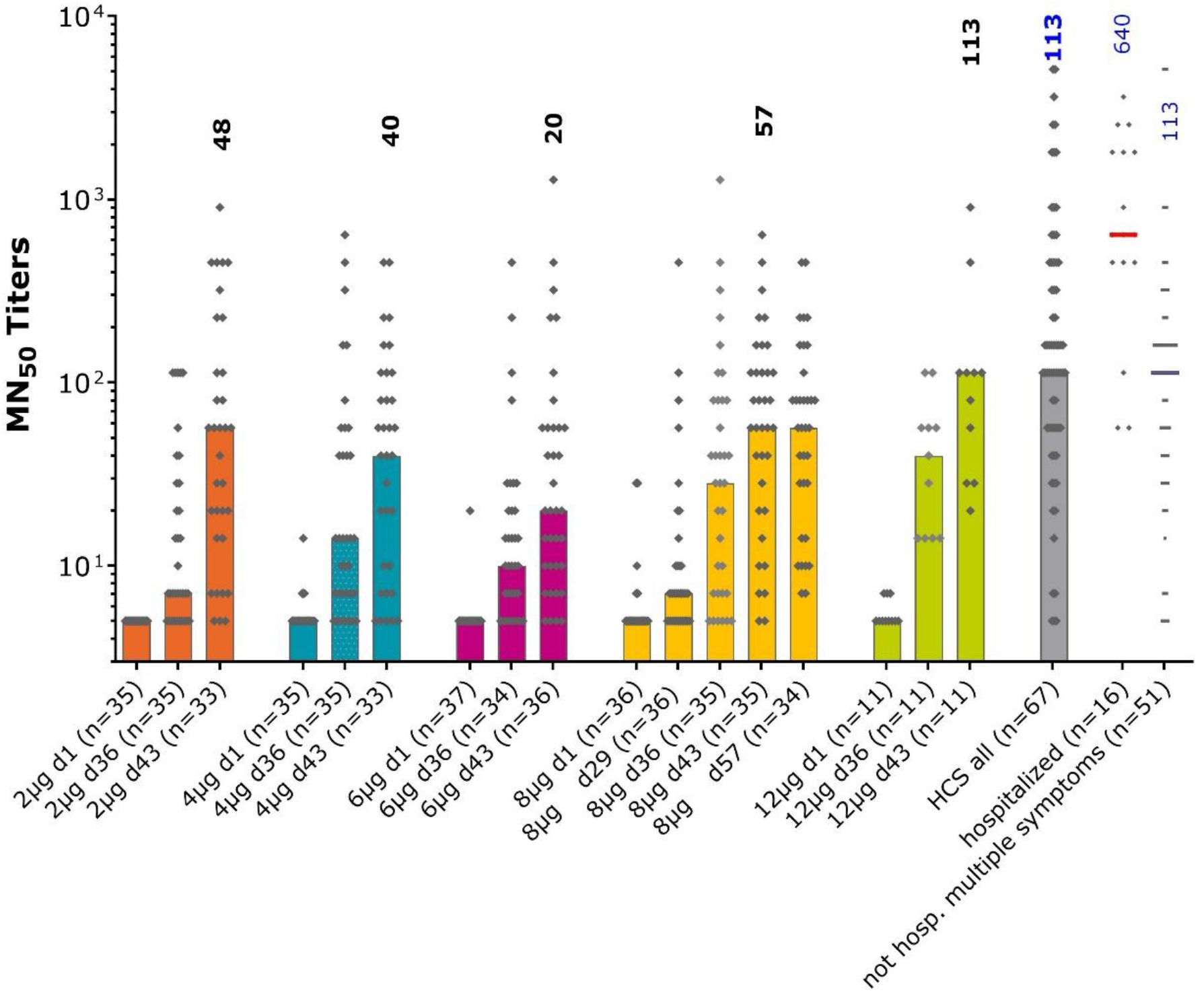
Anti-SARS-CoV-2 virus neutralizing titers in the different study groups and convalescent sera measured by microneutralization. Bars show median values per group at each study timepoint, individual GMT values for each sample shown as diamonds. Numbers show median MN_50_ values at Day 43, two weeks after the second vaccination, for each group and in the convalescent sera.

In subjects **Figure 3** shows the immune response as ELISA IgG antibodies against the S protein with low but variable median titers in the baseline samples (placebo group is not shown as there were no changes in median values over the 50 day period shown). At Day 29, four weeks after the first dose, there were small dose-dependent increases with seroconversion rates of 6– 28% across vaccine groups (**Table 5**). On Day 36, 7 days after the second dose, there was a more marked increase in all groups with 49–82% seroconverting. This rate continued to increase to 79–91% at Day 43 when median titers were 1712 (25th–75th percentile: 789–3132), 2205 (1493–3183), 2839 (1232–7002), 3287 (1470–5770), and 5463 (2675–7132), in 2, 4, 6, 8 and 12 μg groups, respectively. Samples in the 8 μg group from Day 57, four weeks after the second dose, show a small decline in IgG median titers to 1825 (879–2834), but overall these persisted above baseline and the Day 29 (pre-second dose) values. Notably, the value at Day 43 in the 12 μg group was comparable to the median titer of 6381 (5400–12432) in the group of 67 convalescent sera.

**Table 5.** Seroconversion rates in each group at each time-point, n/N (%)

| **Table 5.** Seroconversion rates in each group at each time-point, n/N (%) |  |  |  |  |  |  |
| --- | --- | --- | --- | --- | --- | --- |
|  | **Fold-increase** | **2 µg** | **4 µg** | **6 µg** | **8 µg** | **12 µg** |
| **S protein IgG** |  |  |  |  |  |  |
| **Day 29** | **≥ 4** | 2/35<br>(6) | 6/36<br>(17) | 6/38<br>(16) | 10/36<br>(28) | 2/11<br>(18) |
| **Day 36** | **≥ 4** | 17/35<br>(49) | 25/34<br>(74) | 26/35<br>(74) | 24/35<br>(69) | 9/11<br>(82) |
| **Day 43** | **≥ 4** | 26/33<br>(79) | 27/33<br>(82) | 26/37<br>(70) | 28/35<br>(80) | 10/11<br>(91) |
| **RBD IgG** |  |  |  |  |  |  |
| **Day 36** | **≥ 4** | 6/35<br>(17) | 15/34<br>(44) | 12/34<br>(35) | 23/35<br>(66) | 6/11<br>(55) |
| **Day 43** | **≥ 4** | 13/33<br>(39) | 26/33<br>(79) | 25/36<br>(69) | 29/35<br>(83) | 10/11<br>(91) |
| **Virus neutralizing titers** |  |  |  |  |  |  |
| **Day 36** | **≥ 4** | 12/35<br>(34) | 13/35<br>(37) | 11/34<br>(32) | 21/35<br>(60) | 7/11<br>(64) |
| **Day 43** | **≥ 4** | 24/33<br>(73) | 21/32<br>(66) | 20/36<br>(56) | 27/35<br>(77) | 11/11<br>(100) |

When IgG antibody titers against S protein RBD were assessed (**Figure 4**), an evident response in the 8 μg group data (only group with a full data set in this interim analysis) was shown at Day 29 after one dose. The marked increases in titers 7 days (Day 36) after the second dose were also observed when seroconversion rates were 17–66% (**Table 5**). There was a further increase by Day 43 when the seroconversion rates were 83% and 91% in the 8 and 12 μg groups with median titers of 1240 (349–2952) and 1007 (678–3141), respectively, which were comparable to the median of 1448 (726–5391) observed in convalescent sera.

These observations of IgG antibody responses to S protein and RBD translated into SARS-CoV-2 viral neutralizing activity, as shown in **Figure 5**. This response was less obviously dose-dependent from the available samples, but across the groups 34–64% had seroconverted at Day 36 (7 days after the second dose) from baseline. This rate continued to increase to Day 43 when 100% of the 12 μg group (n = 11) had seroconverted. The median neutralizing titer in this group (113 MNT_50_ [28.3–113]) on Day 43 was the same as that observed in convalescent sera (113 MNT_50_ [56.6–453]). In contrast to S protein IgG levels, the 8 μg group median neutralizing titers at Day 43 (56.5 MNT_50_ [20.0–113]) were generally maintained to Day 57 (56.6 MNT_50_ [24.8–125]).

Immunogenicity of CVnCoV in subjects known to be SARS-CoV-2 seropositive at baseline is presented in **Figure 6**. After vaccination with either with 2µg or 4µg of CVnCoV, a rapid increase in antibody titers is shown within one week post first vaccination. This was observed for both binding and neutralizing antibodies. The effect of a second dose was less pronounced; antibody titers remained stable at least up to day 57.

**Figure 6.**
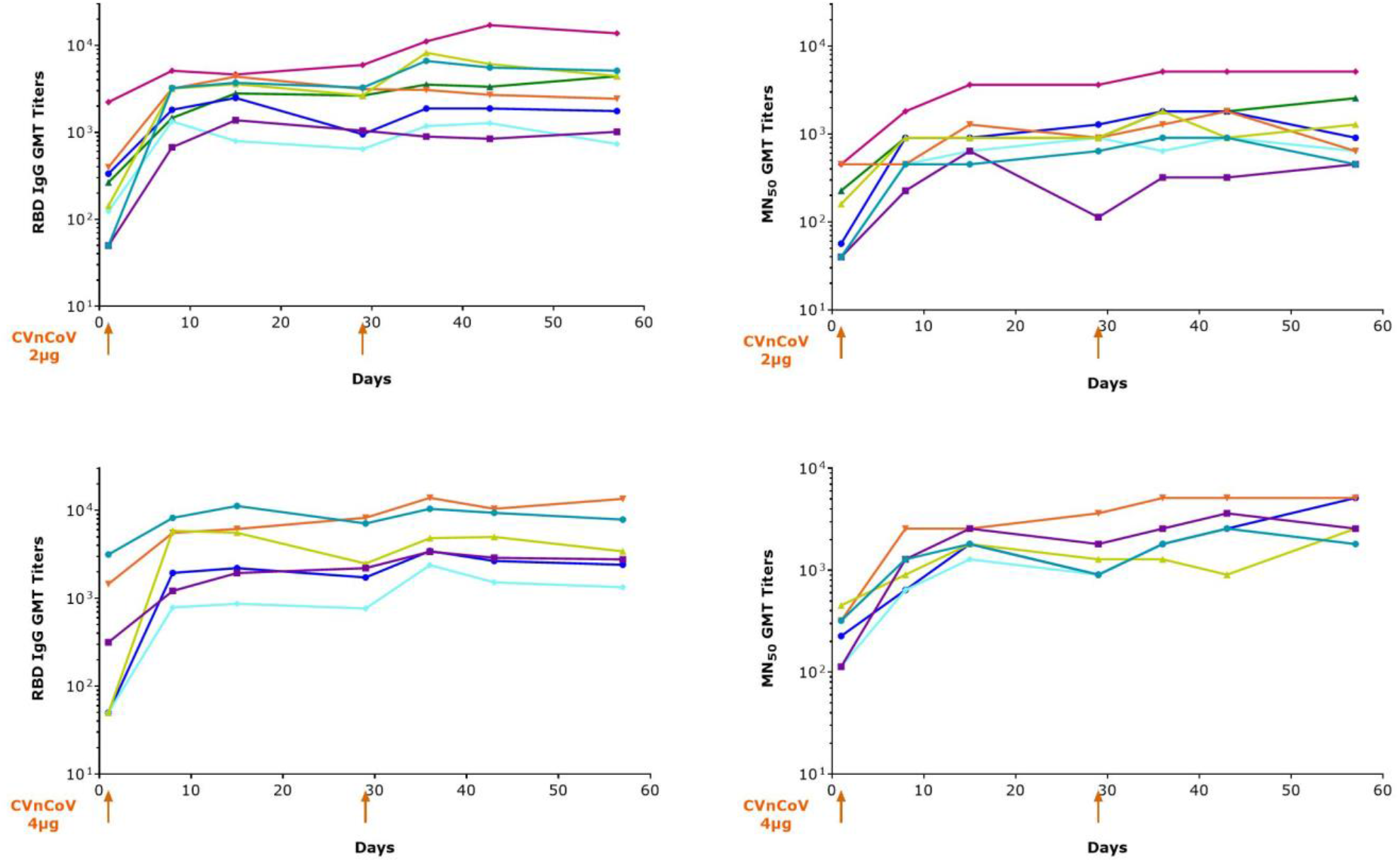
Boosting of antibody responses in seropositive subjects. Seropositve subjects were vaccinated either with 2µg (upper panel) or 4µg (lower panel) of CVnCoV on day 1 and 29. RBD binding antibodies as well as SARS-CoV-2 antibodies were analyzed on multiple time points. Lines show individual subjects in both dose groups.

The analysis of the ratio between binding and neutralizing antibodies (VNT to ELISA RBD and ELISA Spike) was performed for the the selected dose 12 mcg and is presented in **Table 6**. At Day 43 in the study (i.e. after two doses of CVnCoV) the corresponding median of these ratios are comparable of those measured in the convalescent sera (HCS = Human Convalescent Sera) (VNT to ELISA Spike: 0.0218 12 mcg, 0.0213 HCS & VNT to ELISA RBD: 0.0806 12 mcg, 0.0832 HCS).

**Table 6.** Ratio of neutralizing to binding antibodies in vaccinated and convalescent subjects

|  |  |  |
| --- | --- | --- |
| **Dose** | **VNTs/RBD IgG** | **VNT/Spike IgG** |
| **12µg** | 0.0806 | 0.0218 |
| **HCS** | 0.0832 | 0.0213 |

**Table 7.** Demographic and Baseline Characteristics - Safety Population Previously SARS-nCoV-2 seropositives

| Variable | Category | Statistic | Cohort 1<br>(2 ug) | Cohort 2<br>(4 ug) |
| --- | --- | --- | --- | --- |
|  |  |  | (N=8) | (N=8) |
| Age | (Years) | Mean (SD) | 39.6 (12.58) | 39.1 (12.45) |
|  |  | Median | 41 | 41 |
|  |  | Min, Max | (20, 54) | (19, 54) |
| Gender | Male | n (%) | 5 (63%) | 6 (75%) |
|  | Female | n (%) | 3 (38%) | 2 (25%) |
| BMI | (kg/m<sup>2</sup>) | Mean (SD) | 23.8 (2.076) | 25.4 (3.464) |
|  |  | Median | 24 | 26 |
|  |  | Min, Max | (21, 28) | (20, 30) |
| RACE | Asian | n (%) | 0 (0%) | 0 (0%) |
|  | White | n (%) | 8 (100%) | 8 (100%) |
| Ethnicity | Not Hispanic or Latino | n (%) | 8 (100%) | 8 (100%) |

**Table 8.**
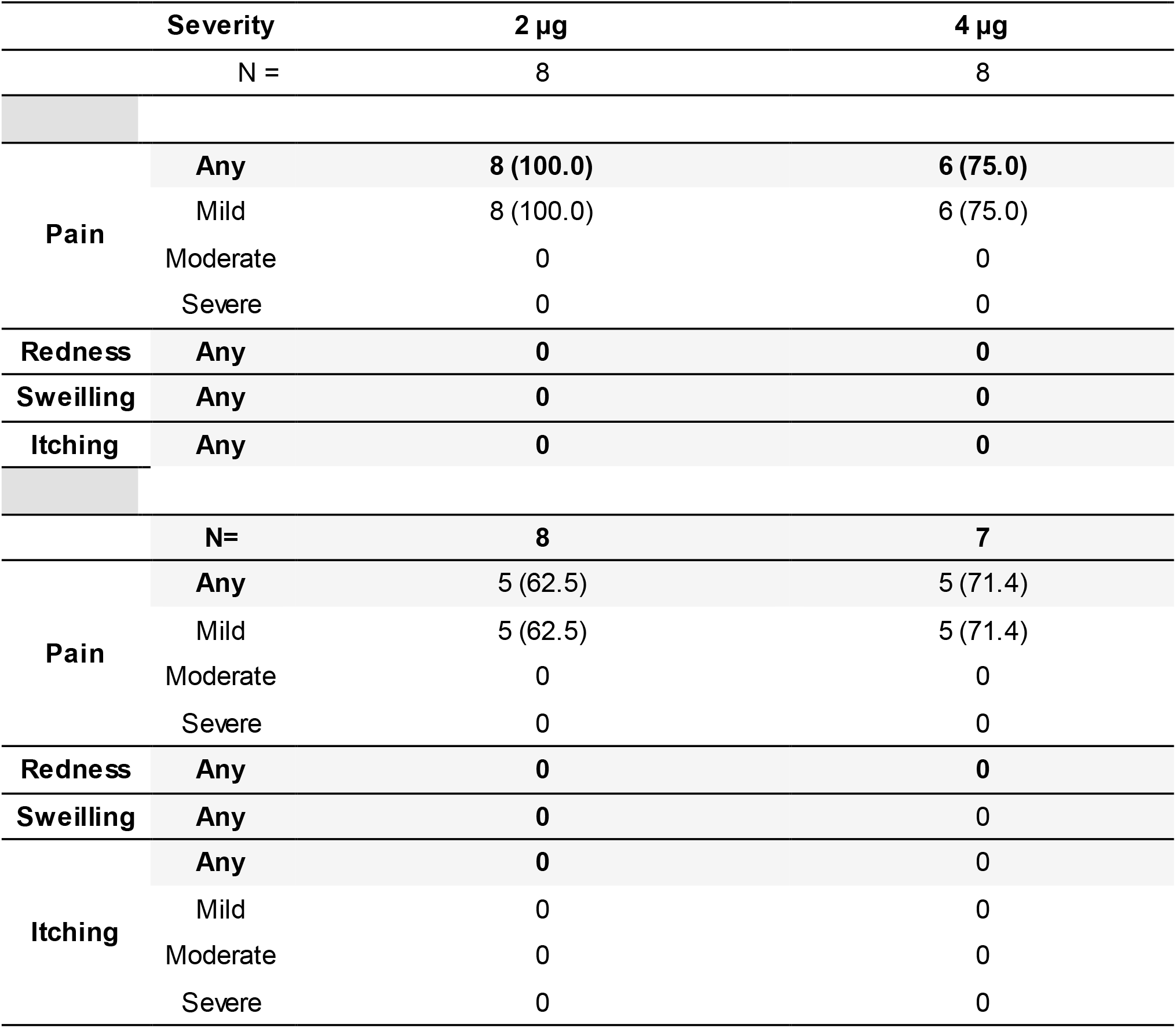
Solicited local AEs with severity after one dose

**Table 9a.** Solicited systemic AEs with severity within 7 days after the first dose

| **Severity** |  | **2 µg** | **4 µg** |
| --- | --- | --- | --- |
| N = |  | 8 | 8 |
| **Fever** | **Any** | **0** | **2 (25.0)** |
|  | Mild | 0 | 1 (12.5) |
|  | Moderate | 0 | 1 (12.5) |
|  | Severe | 0 | 0 |
| **Headache** | **Any** | **3 (37.5)** | **5 (62.5)** |
|  | Mild | 3 (37.5) | 5 (62.5) |
|  | Moderate | 0 | 0 |
|  | Severe | 0 | 0 |
| **Fatigue** | **Any** | **5 (62.5)** | **5 (62.5)** |
|  | Mild | 4 (50.0) | 3 (37.5) |
|  | Moderate | 1 (12.5) | 2 (25.0) |
|  | Severe | 0 | 0 |
| **Chills** | **Any** | **0** | **3 (37.5)** |
|  | Mild | 0 | 2 (25.0) |
|  | Moderate | 0 | 1 (12.5) |
|  | Severe | 0 | 0 |
| **Myalgia** | **Any** | **2 (25.0)** | **4 (50.0)** |
|  | Mild | 2 (25.0) | 3 (37.5) |
|  | Moderate | 0 | 1 (12.5) |
|  | Severe | 0 | 0 |
| **Arthralgia** | **Any** | **0** | **2 (25.0)** |
|  | Mild | 0 | 2 (25.0) |
|  | Moderate | 0 | 0 |
|  | Severe | 0 | 0 |
| **Nausea/**<br>**Vomiting** | **Any** | **1 (12.5)** | **1 (12.5)** |
|  | Mild | 1 (12.5) | 1 (12.5) |
|  | Moderate | 0 | 0 |
|  | Severe | 0 | 0 |
| **Diarrhea** | **Any** | **1 (12.5)** | **1 (12.5)** |
|  | Mild | 1 (12.5) | 1 (12.5) |
|  | Moderate | 0 | 0 |
|  | Severe | 0 | 0 |

**Table 9b.** Solicited systemic AEs with severity within 7 days after the second dose

| **Severity** |  | **2 µg** | **4 µg** |
| --- | --- | --- | --- |
| N = |  | 8 | 7 |
| **Fever** | **Any** | **0** | **0** |
|  | Mild | 0 | 0 |
|  | Moderate | 0 | 0 |
|  | Severe | 0 | 0 |
| **Headache** | **Any** | **3 (37.5)** | **4 (57.2)** |
|  | Mild | 2 (25.0) | 3 (42.9) |
|  | Moderate | 1 (12.5) | 0 |
|  | Severe | 0 | 1 (14.3) |
| **Fatigue** | **Any** | **4 (50.0)** | **4 (57.2)** |
|  | Mild | 3 (37.5) | 3 (42.9) |
|  | Moderate | 1 (12.5) | 0 |
|  | Severe | 0 | 1 (14.3) |
| **Chills** | **Any** | **1 (12.5)** | **0** |
|  | Mild | 1 (12.5) | 0 |
|  | Moderate | 0 | 0 |
|  | Severe | 0 | 0 |
| **Myalgia** | **Any** | **2 (25.0)** | **1 (14.3)** |
|  | Mild | 2 (25.0) | 1 (14.3) |
|  | Moderate | 0 | 0 |
|  | Severe | 0 | 0 |
| **Arthralgia** | **Any** | **0** | **3 (42.9)** |
|  | Mild | 0 | 3 (42.9) |
|  | Moderate | 0 | 0 |
|  | Severe | 0 | 0 |
| **Nausea/**<br>**Vomiting** | **Any** | **0** | **1 (14.3)** |
|  | Mild | 0 | 1 (14.3) |
|  | Moderate | 0 | 0 |
|  | Severe | 0 | 0 |
| **Diarrhea** | **Any** | **0** | **0** |
|  | Mild | 0 | 0 |
|  | Moderate | 0 | 0 |
|  | Severe | 0 | 0 |

## DISCUSSION

In this ongoing phase 1 clinical trial, we are investigating increasing dose levels, from 2 to 12 µg of the mRNA vaccine candidate CVnCoV in a two-dose schedule in healthy adults from 18 to 60 years of age. We present here an interim report of data obtained so far. We found that the vaccine was safe and showed an acceptable reactogenicity profile at all levels, including a subset of subjects who were known to be SARS-CoV-2 seropositive at baseline. Compliance with the vaccination schedule was high, with only four subjects not receiving the second dose due to AEs. There were no vaccine-related SAEs and although the incidence and severity of solicited adverse reactions appears to increase at increasing dose levels, reactogenicity was not dose-limiting, and higher dose levels (16 μg and 20 μg) are currently being investigated. Observed local reactions were almost exclusively cases of transient mild to moderate injection site pain; of the 415 total administered doses of CVnCoV at any dosage only three resulted in transient severe local pain, all after the first dose (one each in the 2, 4 and 8 µg groups). The frequency and severity of solicited systemic AEs increased with dosage level and were generally more frequent and of higher intensity after the second dose than the first, as has been observed with other mRNA SARS-CoV-2 vaccine candidates [20–22]. These systemic AEs mainly consisted of transient mild or moderate headache and fatigue, and to a lesser extent myalgia and chills. Overall, fever was observed less frequently. Severe solicited AEs usually decreased or disappeared rapidly, mostly within 24–48 hours of onset. The reactogenicity profile, with limited presence of fever but symptoms like fatigue, headache and chills, is probably associated with the postulated mechanism of action and induction of an innate immune response mediated by interferon and other immune-stimulatory cytokines.

Th1 cytokines are important for development of T cell responses, CD4 T cell help is required for good induction of memory B cells. Moreover, such a T-helper cell type 1 (Th1) biased immune response is desirable for the development of a SARS-CoV-2 prophylactic vaccine, due to the hypothetical concern for immune-mediated disease enhancement observed in preclinical studies for other coronaviruses. IFN type 1 signaling has been also described in COVID patients as a critical pathway to control disease [23,24].

All investigated dosages elicited an immune response against SARS-CoV-2. In this interim report, induction of an adaptive humoral immune response was demonstrated by the increase in neutralizing antibodies, with 56–77% of participants achieving VNT seroconversion two weeks after two doses of 2–8 μg, and 100% seroconverting two weeks after two 12 μg doses. This neutralizing activity was associated with marked S protein-specific and RBD-specific IgG antibody responses. S protein IgG and VNT responses were detectable after the first vaccination, but all markedly increased within 7 days of the second vaccination indicating efficient priming by the first dose. In the 8 μg group the immune response persisted up to at least Day 57, the last time-point with data currently available.

Since an imbalance between binding versus neutralizing antibodies could hypothetically lead to immune-mediated disease enhancement, we investigated the ratio neutralizing/binding antibodies both for the S Spike protein as for RBD in the study participants. We calculated such ratios on an individual basis for vaccinated study participants as well as for convalescent patients (Table 6). We observed that the ratio post vaccination is very similar as the ratio measured in sera from convalescent patients after natural infection. This observation is also in line with the mechanism of action of CVnCoV which mimics natural immune response to RNA viruses.

Data in SARS-CoV-2 seropositive subjects show CVnCoV is well tolerated, supporting further use of the candidate vaccine in future clinical development and a broader population without the need for previous testing for serostatus and excluding seropositive subjects. Furthermore, the data show that good memory responses are induced by the natural infection, since even in subjects with low antibody titers at baseline, low doses of CVnCoV (either 2µg or 4µg) were able to expand antibody titers by more than a factor 10 within one week after the first vaccination. This supports the role of memory cells to provide a long term protection to SARS-CoV-2.

Our recently shared preclinical data in a hamster model showed good priming followed vaccination with low dose of CVnCoV or single vaccination and rapid boosting of neutralizing antibodies followed virus challenge. This was comparable to the groups vaccinated with two doses of vaccine [9]. The available preclinical and clinical data are indicative of a functional immune response mimicking the natural responses to infection, including a potent induction of memory. More analyses on T- and B-cell memory responses are currently ongoing in this clinical study, as well as in preclinical studies with CVnCoV and will further inform the unique mechanism of action of this mRNA vaccine candidate.

The study is ongoing with additional testing of higher doses (16 μg and 20 μg) to investigate the boundaries of the safety window and completion of the assessments of the present groups, with follow up foreseen until at least 1-year post vaccination. Based on the need to adequately balance an acceptable reactogenicity profile with a strong immune response in the range of convalescent sera (including 100% seroconversion for VNT), the 12 μg dosage has been selected for further investigation in phase 2 and 3 studies, most notably in a phase 2b/3 study to assess efficacy and safety in 36,500 participants that will begin in the coming weeks.

## Data Availability

No additional data are referred to

