## Supplementary Material for "Phase 1 Assessment of the Safety and Immunogenicity of an mRNA- Lipid Nanoparticle Vaccine Candidate Against SARS-CoV-2 in Human Volunteers"

| Intensity Grading for Solicited Local Adverse Events | | |
|---|---|---|
| **AE** | **Grade** | **Definition** |
| Pain at injection site | 0 | Absent |
| | 1 | Does not interfere with activity |
| | 2 | Interferes with activity and/or repeated use of non-narcotic pain reliever > 24 hours |
| | 3 | Prevents daily activity and/or repeated use of narcotic pain reliever |
| Redness | 0 | ≤ 2.5 cm |
| | 1 | 2.5 – 5 cm |
| | 2 | 5.1 – 10 cm |
| | 3 | >10 cm |
| Swelling | 0 | ≤ 2.5 cm |
| | 1 | 2.5 – 5 cm and does not interfere with activity |
| | 2 | 5.1 – 10 cm or interferes with activity |
| | 3 | > 10 cm or prevents daily activity |
| Itching | 0 | Absent |
| | 1 | Mild, no interference with normal activity |
| | 2 | Moderate, some interference with normal activity |
| | 3 | Significant, prevents normal activity |

| **Intensity Grading for Solicited Systemic Adverse Events** | | |
|---|---|---|
| **Adverse Event** | **Grade** | **Definition** |
| Fever | 0 | <38°C |
| | 1 | ≥38 – 38.4°C |
| | 2 | ≥38.5 – 38.9°C |
| | 3 | ≥39°C |
| Headache | 0 | Absent |
| | 1 | Mild, no interference with normal activity |
| | 2 | Moderate, some interference with normal activity and/or repeated use of non-narcotic pain reliever >24 hours |
| | 3 | Significant; any use of narcotic pain reliever and/or prevents daily activity |
| Fatigue | 0 | Absent |
| | 1 | Mild, no interference with normal activity |
| | 2 | Moderate, some interference with normal activity |
| | 3 | Significant, prevents normal activity |
| Chills | 0 | Absent |
| | 1 | Mild, no interference with normal activity |
| | 2 | Moderate, some interference with normal activity |
| | 3 | Significant, prevents normal activity |
| Myalgia | 0 | Absent |
| | 1 | Mild, no interference with normal activity |
| | 2 | Moderate, some interference with normal activity |
| | 3 | Significant, prevents normal activity |
| Arthralgia | 0 | Absent |
| | 1 | Mild, no interference with normal activity |
| | 2 | Moderate, some interference with normal activity |
| | 3 | Significant, prevents normal activity |
| Nausea/ Vomiting | 0 | Absent |
| | 1 | Mild, no interference with activity and/or 1 – 2 episodes/ 24 hours |
| | 2 | Moderate, some interference with activity and/or >2 episodes/ 24 hours |
| | 3 | Significant, prevents daily activity, requires outpatient i.v. hydration |
| Diarrhea | 0 | Absent |
| | 1 | 2 – 3 loose stools or <400 g/24 hours |
| | 2 | 4 – 5 stools or 400 – 800 g/24 hours |
| | 3 | 6 or more watery stools or >800 g/24 hours or requires outpatient i.v. hydration |

i.v.= Intravenous
